# Risk of Ocular Adverse Events with Taxane Based Chemotherapy

**DOI:** 10.1101/2022.02.15.22270863

**Authors:** Mohit Sodhi, Sonia N. Yeung, David Maberley, Frederick Mikelberg, Mahyar Etminan

## Abstract

**OBJECTIVE:** Taxane based chemotherapy agents such as docetaxel and paclitaxel are used for a wide range of cancers. Although much has been published on adverse events related to taxanes, data on ocular outcomes with these very important drugs is scant. We sought to quantify the risk of three ocular adverse events: 1) epiphora, 2) cystoid macular edema (CME) and 3) optic neuropathy with taxane based chemotherapy agents by undertaking a large pharmacoepidemiologic study.

**DESIGN:** New users, retrospective cohort study design

**SUBJECTS, PARTICIPANTS, CONTROLS:** We created a cohort of new users of women matched by age (within one year) on docetaxel or paclitaxel and new users of tamoxifen as controls. Study members were followed to the first incidence of the three outcomes.

**METHODS:** Descriptive statistics were used to examine demographics of the exposed and unexposed groups. A Cox model was constructed to compute crude and adjusted hazard ratios. Potential confounders for each outcome were adjusted for.

**MAIN OUTCOME MEASURES:** First diagnosis of 1) epiphora, 2) cystoid macular edema (CME) or 3) optic neuropathy ascertained using international classification for diseases 9^th^ or 10^th^ division codes.

**RESULTS:** For the epiphora analysis there were 1,824 taxane users (PTX or DTX) and 16,395 tamoxifen users. The crude HR for this association was 7.32 (95%CI: 4.25-16.61) and adjusted HR was 5.15 (95% CI:2.79-9.54). For the CME analysis there were 1,909 taxane user and 16,524 tamoxifen users. The crude HR for CME comparing taxane users to tamoxifen users was 1.37 (95% CI: 0.72-2.60) and adjusted HR was 1.33 (95% CI: 0.70-2.53). For optic neuropathy there were 1,913 taxane users and 16,566 tamoxifen users. The crude HR was 4.43 (95% CI:1.10-17.82) and the adjusted HR was 4.44 (95% CI:1.04-18.87). When we restricted our analysis to only cases of toxic optic neuropathy the aged-matched HR remained elevated (HR=7.24, 95% CI: 1.13-46.36).

**CONCLUSION:** In a cohort of women who were using taxane chemotherapy agents, there was an elevated risk for epiphora, optic neuropathy and a less precise increase in risk for cystoid macular edema. Ophthalmologists and oncologists should be aware of these adverse events in women with breast cancer who use these drugs.

## INTRODUCTION

Chemotherapy remains the mainstay therapy for most cancers including breast cancer, lung cancer, genitourinary cancers and head and neck cancers^1^. Although much is known about adverse events related to taxanes such as docetaxel (DTX) and paclitaxel (PTX), information on the frequency of ocular adverse events including their incidence is scant. A number of ocular adverse events secondary to PTX or DTX have been published which have mainly focused on 1) epiphora^2-4^, 2) cystoid macular edema (CME^5,6^), and 3) optic neuropathy^7^. Epiphora is a significantly debilitating condition that may be irreversible and greatly affects quality of life, in particular, outdoor activities^8^ and research has shown that if treatable, correction of epiphora can vastly improve overall health and quality of life^9^. CME and optic neuropathy are also serious adverse events, and their prompt diagnosis and treatment can affect patient outcomes. Most of the data on these three adverse events with PTX and DTX have been mainly reported as case reports which cannot quantify a true incidence or provide an effect size as to the magnitude of these events.

In 2020 more than 2 million women were diagnosed with breast cancer around the world^10^. Approximately 60% of women with breast cancer are diagnosed with invasive breast cancer. which usually require chemotherapy that includes PTX or DTX^11^. Therefore, ophthalmologists and oncologists need to be familiar with the frequency and type of ocular adverse advents secondary to taxane chemotherapy. Recognizing these ocular adverse events early will potentially prevent complications and improve patient outcomes. Therefore, we sought to quantify the risk of these adverse events by undertaking a large pharmacoepidemiologic study.

## METHODS

### Data Sources

We utilized the PharMetrics Plus database (IQVIA, Durham, NC) as the primary data source for the study. The PharMetrics database is a private health claims database from the United States of America that captures health information of more than 150 million enrollees in the USA^12^. The data captured include: demographics (including the age and sex of patients), physician diagnosis (through the international classification for disease ninth or tenth editions clinical modification (ICD-9 CM or ICD-10) codes, and prescription drug information (drug name, day supply, and dose). We had access to a random sample of this database from 2006 to 2020 on 16,424,887 subjects with at least one year of enrollment. All analyses were performed using SAS version 9.4. Approval for the study was obtained by the University of British Columbia ‘s Clinical Research Ethics Board.

### Cohort Description

We created a cohort of women who were new users of DTX and PTX, the two most prescribed taxanes in women. New users were defined as those who were not using either drug or did not have any of the study outcomes in the year prior to cohort entry. We also included women who were new users of tamoxifen for the treatment of estrogen positive breast cancer who were not using PTX, DTX or fluorouracil, another chemotherapy drug known to increase the risk of epiphora. Tamoxifen was chosen as a reference drug because it is used for similar indications (breast cancer) as taxanes, and it has shown to increase the risk of CME^13^ and optic neuropathy^14^, although it has not been associated with epiphora. However, since the mechanism of action of taxanes and tamoxifen are different, we decided to use tamoxifen as a comparator against taxanes. Cohort members were followed to the first of the following events: 1) epiphora which also included stenosis of lacrimal canaliculi and obstruction of nasolacrimal ducts (ICD-9 codes: 375.20, 375.53,375.55 and ICD-10 codes: H04.54 or H04), 2) cystoid macular edema (ICD-9 codes: 362.53, 362.83 and ICD-10 codes: H35.351, H35.352, H35.353, H35.81 and 3) optic neuropathy (ICD-9 codes: 377.34, 377.41 and ICD-10 codes: H47.013, H47.012, H47.011, H47.019, H46.3). Since current data show that taxanes might cause both ischemic optic neuropathy and toxic optic neuropathy^7^ both types were included but a separate analysis for both was also undertaken. We also censored subjects who switched from a taxane to tamoxifen or vice versa.

### Statistical analysis

We used descriptive statistics to examine demographics and covariate distribution between the exposed groups (PTX or DTX) and controls (tamoxifen users). Our study was matched by age and sex (all female cohort) by design (the two main confounders for these study questions). We further constructed a Cox regression model to compute crude and adjusted hazard ratios (HRs) for the three outcomes comparing DTX or PTX with tamoxifen users. For the model looking at epiphora, we adjusted for the following covariates (mainly risk factors) keratoconjuctivitis, dry eyes syndrome, keratitis, blepharitis, and lid malposition. For the model looking at CME, we adjusted for previous cataracts and diabetes. For the model examining optic neuropathy we also adjusted for hypertension, coronary artery disease, diabetes, and number of ophthalmologist visits. We computed incident rates and adjusted hazard ratios for all three outcomes.

## RESULTS

For the epiphora analysis there were 1,824 taxane users (PTX or DTX) and 16,395 tamoxifen users. The incidence of epiphora among taxane users was 55.6/10,000 person years vs. 7.9/10,000 years for tamoxifen users. The crude HR for this association was 7.32 (95% CI: 4.25-16.61) and adjusted HR of 5.15 (95% CI: 2.79-9.54). For the CME analysis there were 1,909 taxane user and 16,524 tamoxifen users. The incidence of CME among taxane users was 34.8/10,000 vs. 16.8/10,000 users in the tamoxifen users. The crude HR for CME comparing taxane users to tamoxifen users was 1.37 (95%CI :0.72-2.60) and the adjusted HR was 1.33 (95% CI: 0.70-2.53). For optic neuropathy there were 1,913 taxane users and 16,566 tamoxifen users. The incidence of optic neuropathy for taxane users was 10.6/10,000 1.2/10,000 for tamoxifen users. The crude HR was 4.43 (95% CI: 1.10-17.82) and the adjusted HR was 4.44 (95% CI: 1.04-18.87). When we restricted our analysis to only cases of toxic optic neuropathy the aged-matched HR remained elevated (HR=7.24, 95% CI: 1.13-46.36).

## DISCUSSION

The results of our study suggests that taxane based chemotherapy agents can increase in the risk of epiphora and optic neuropathy. There was also an elevated risk with CME, but results were not precise and confidence intervals were wide (HR=1.33, 95% CI :0.70-2.53). Our study results are consistent with published case-reports that have suggested an increase in the risk of these three important ocular adverse events^2,5,8^.

There is a paucity of evidence surrounding the potential pathophysiological mechanism of taxane induced optic neuropathy. It has been hypothesized that the systemically administered taxanes can cause neurotoxic or ischemic damage to the retinal ganglion cells^7^. Other research has shown that taxanes can induce maladaptive electrophysiological changes within the retina and optic nerve which were attributed to vasospasm or vascular dysregulation^15^. This hypothesis is supported by findings from case studies such that the patient presentation of taxane induced optic neuropathy resembles that of interferon alpha therapy, which is also believed to be caused by ischemia and vascular dysregulation^7^. Furthermore, since taxanes are known to be neurotoxic and can cause peripheral neuropathy, it is possible that the optic nerve is damaged via a similar mechanism^16^.

The proposed mechanism for taxane induced epiphora is canalicular stenosis. The drug is secreted in the tear film, and chronic inflammation of the canaliculi as these tears are drained through the nasolacrimal system results in scarring and narrowing of the passageway.^2, 17^. However, epiphora has also been shown to occur in the absence of canalicular stenosis and is believed to be secondary to dry eye disease caused by taxane based chemotherapy^2^. The use of artificial tears may not only serve to dilute and clear the drug from the ocular surface through the nasolacrimal system, but may treat any related dry eye disease. Early recognition and discontinuation of the drug if possible, may prevent surgical intervention^17^. Alternatively, silicone intubation may be an option to manage canalicular stenosis in the early stages. However, delayed diagnosis and management may lead to obstruction along the nasolacrimal system necessitating more complex surgery^17^.

It has been postulated that CME can result of a disruption of the retinal-blood barrier and fluid accumulation in intracellular spaces^18^. Some have also proposed that taxanes can cause retinal cell toxicity by due to taxanes inherent mechanism of inhibiting microtubular reorganization^18^.

There are more than 2 million women suffering from breast cancer worldwide^10^ and more than 60%^11^ of these women are likely undergoing taxane based chemotherapy. There is projected to be nearly 2.9 billion^19^ USD in sales of taxanes, indicating that utilization of these drugs will greatly increase over the next few years for both breast and other types of cancers. Therefore, the ocular adverse events studied will become more significant in the near future.

Our study has strengths and limitations. This was the first and largest study of women who used a taxane (with aged-matched controls using tamoxifen) that examined three important ocular adverse events. The strength of our study is use of a comparator group and adjustment for other confounder or risk factors. Although most taxanes in women are used for breast cancer, it is possible that they were used for other types of cancer. However, we don ‘t believe different types of cancers would alter the risk of these three ocular adverse events. As with all pharmacoepidemiologic studies that use claims data we did not have access to patients ‘ charts for outcome validation.

In a cohort of women who were using taxane chemotherapy agents, there was an elevated risk for epiphora, optic neuropathy and a less precise increase in risk for cystoid macular edema. Ophthalmologists and oncologists should be aware of these adverse events in women with breast cancer who use these drugs. Early identification of such events may help in treatment of complications and lead to better outcomes and quality of life. Future studies should confirm these results.

## Data Availability

Data is available upon reasonable request as contained in the manuscript

---

**Table 1.** Characteristics of Tamoxifen and Docetaxel/Paclitaxel Users

|  | **Tamoxifen** | **Docetaxel/Paclitaxel** |
| --- | --- | --- |
| **Epiphora** |  |  |
| Number | 16,395 | 1,824 |
| Age (mean ± sd) | 54.6 ± 12.8 | 62.1 ± 12.7 |
| Follow-up | 3.5 ± 3.1 | 2.0 ± 1.9 |
| Events | 45 | 20 |
| Rate | 7.9 / 10,000 person years | 55.6 / 10,000 person years |
| Keratoconjunctivitis (%) | 0.05 | 0.11 |
| Dry eyes (%) | 0.29 | 2.25 |
| Keratitis (%) | 2.07 | 3.95 |
| Blepharitis (%) | 2.16 | 4.61 |
| Lid malposition (%) | 0.55 | 1.35 |
| **Cystoid Macular Edema** |  |  |
| Number | 16,524 | 1,909 |
| Age (mean ± sd) | 54.6 ± 12.8 | 62.5 ± 12.8 |
| Follow-up | 3.5 ± 3.1 | 2.0 ± 1.9 |
| Events | 96 | 13 |
| Rate | 16.8 / 10,000 person years | 34.8 / 10,000 person years |
| Cataract (%) | 0.6 | 2.1 |
| Diabetes (%) | 8.0 | 14.3 |
| **Optic Neuropathy** |  |  |
| Number | 16,395 | 1,824 |
| Age (mean ± sd) | 54.6 ± 12.8 | 62.5 ± 12.8 |
| Follow-up | 3.5 ± 3.1 | 2.0 ± 1.9 |
| Events | 7 | 4 |
| Rate | 1.2 / 10,000 person years | 10.6 / 10,000 person years |
| Hypertension (%) | 6.0 | 38.2 |
| Diabetes (%) | 9.9 | 17.4 |
| Coronary artery disease (%) | 5.7 | 18.6 |
| Ophthalmology visits (mean ± sd) | 0.65 ± 2.11 | 0.92 ± 2.88 |

**Table 2.** Hazard ratios for cases of epiphora, cystoid macular edema, and optic neuropathy with exposure to Tamoxifen and Docetaxel/Paclitaxel

|  | **Crude**<br>**HR** | **Crude 95%**<br>**CI** | **Adjusted**<br>**HR** | **Adjusted 95%**<br>**CI** |
| --- | --- | --- | --- | --- |
| **Epiphora** |  |  |  |  |
| Tamoxifen | 1 | Reference | 1.00 | Reference |
| Docetaxel / Paclitaxel | 5.55 | 2.99-10.28 | 5.15 | 2.79-9.54 |
| **Cystoid Macular Edema** |  |  |  |  |
| Tamoxifen | 1 | Reference | 1.00 | Reference |
| Docetaxel / Paclitaxel | 1.37 | 0.72-2.60 | 1.33 | 0.70-2.53 |
| **Optic Neuropathy** |  |  |  |  |
| Tamoxifen | 1 | Reference | 1.00 | Reference |
| Docetaxel / Paclitaxel | 4.33 | 1.10-17.82 | 4.44 | 1.04-18.87 |

## Notes

**Conflicts of interest:** None of the authors have any conflicts to report.

### Competing Interest Statement

The authors have declared no competing interest.

### Funding Statement

The study did not receive any funding

### Author Declarations

Ethics committee of the University of British Columbia gave ethics approval for this work

